# Germline genetic variants and epilepsy surgery response: individual-participant pooled analysis of 269 patients

**DOI:** 10.64898/2026.03.10.26347961

**Authors:** Alina Ivaniuk, Sunanjay Bajaj, Christian M Boßelmann, Hyun Yong Koh, Elia Pestana-Knight, Xiaoming Zhang, William Bingaman, Imad Najm, Manish Shah, Nitin Tandon, Gretchen Von Allmen, Samden D. Lhatoo, William Tatum, Brin Freund, Kai J. Miller, Elaine Wirrell, Anthony Fine, Jason Coryell, John Schreiber, Scott Perry, Pediatric Epilepsy Research Consortium (PERC) Surgery Workgroup, Pediatric Epilepsy Research Consortium (PERC) Genetics Workgroup, Dennis Lal

**Author notes:** Corresponding Author: Dennis Lal, PhD.

## Abstract

**Background:** Genetic testing is increasingly used in presurgical evaluation, but the yield of resection across germline genetic epilepsies remains uncertain.

**Methods:** We conducted a systematic review of MEDLINE (PubMed) and Scopus and added cases from three institutional cohorts and the Pediatric Epilepsy Research Consortium (PERC) databases, including individuals with pathogenic/likely pathogenic germline variants besides tuberous sclerosis and neurofibromatosis who underwent resection or laser ablation. Etiologies were grouped into biologically informed categories (GATORopathies, vascular, overgrowth, CNVs, channelopathies, synaptopathies, other). Primary outcome was seizure freedom (Engel I) at last follow-up. Group comparisons used Fisher’s exact and Kruskal–Wallis tests (α=0.05; Bonferroni when applicable). Prespecified sensitivity analyses stratified by lesional status, excluded GATORopathies, and restricted to literature-only cases.

**Results:** We included 223 literature cases (64 studies), 35 institutional cases, and 11 PERC cases (n=269). Median follow-up was 24 months (IQR 12–48.0). Seizure freedom was achieved in: vascular disorders 14/19 (73.6%), GATORopathies 79/120 (67.5%), CNVs 18/31 (66.7%), overgrowth 7/13 (53.8%), other 16/25 (41.7%), channelopathies 13/43 (33.3%), and synaptopathies 4/18 (22.2%) (overall p<0.001). Among cases with known imaging, 208/253 (82.2%) had epileptogenic MRI lesions, including 64% of channelopathies and 80% of the synaptopathies. In univariate contrasts (each category vs all others), odds of seizure freedom were higher for vascular disorders (2.66-fold, 95% CI 0.87–9.72) and GATORopathies (2.46-fold, 95% CI 1.46–4.18), and lower for synaptopathies (∼4.2-fold lower, OR 0.24, 95% CI 0.05– 0.78) and channelopathies (∼4.5-fold lower, OR 0.22, 95% CI 0.09–0.48). Direction and magnitude were consistent across prespecified sensitivity analyses (lesional-only, literature-only, exclusion of GATORopathies).

**Conclusions:** Resective surgeries can be effective in germline genetic epilepsies, but outcomes vary by pathway. Disorders with discrete, lesional substrates (GATORopathies, vascular) show the highest likelihood of seizure freedom, whereas channelopathies and synaptopathies, despite the presence of MRI lesions, have substantially lower yields even in lesional cases. Prospective, genotype-aware surgical registries with standardized reporting (EEG, imaging, pathology) and time-to-event outcomes are needed to refine the selection of surgical candidates and quantify seizure and non-seizure-related outcomes.

## Background

In one-third of individuals with focal epilepsy, seizures are not controlled with anti-seizure medications despite the availability of newer agents.^1^ For appropriately selected individuals with medication-resistant epilepsy, epilepsy surgery presents a safe and effective treatment option. Well-defined epileptogenic zone and the presence of a corresponding lesion on brain imaging have been associated with good surgical outcomes across different studies and are thus major determinants of surgical candidacy.^2,3^ With the rising use of genetic testing in epilepsy, genetic diagnosis has become an additional variable in presurgical decision-making and might influence these established surgical prognostic factors.^4–6^

Advances in clinical genetic testing led to expansion of known phenotypic spectra of epilepsies caused by germline genetic variants – variants that affect germline cells and thus are present in all cell of an organism.^7,8^ Genetic etiology thus becomes an increasingly common clinical variable in presurgical assessment of individuals with epilepsy.^9^ For a number of germline genetic epilepsies (GE), focal epilepsy is a main phenotypic feature and thus presents an opportunity for effective surgical treatment – such as for GATOR-related epilepsies caused by variants in *DEPDC5* and *NPRL2*/*NPRL3*.^9^ In contrast, reports on real-world use of genetic testing in presurgical evaluation indicate that revealing genetic etiologies of epilepsy associated with diffuse, multifocal, or generalized epileptogenicity, such as channelopathies and synaptopathies, tends to re-route individuals from the resective surgery candidate pool to palliative approaches, such as neuromodulation and callosotomy.^4,10,11^ Nevertheless, the number of individual case reports and small cohort studies reporting resective surgery across different GE has increased over the past decade. Two prior systematic reviews summarized surgery in genetic epilepsies but were limited for presurgical decision-making. ^5,12^ Stevelink et al. reported ∼50% seizure freedom overall but pooled germline and somatic etiologies and offered limited pathway-specific resolution. ^17^ Cui et al. similarly estimated ∼55% seizure freedom across mixed etiologies, without individual-level, pathway-aware analyses. ^5,12^ Both reviews largely reflected heterogeneous reporting and did not quantify outcomes by genetic pathway. Consequently, clinicians still lack contemporary, genotype-informed estimates to guide candidacy and counseling.

Here, we aim to provide an up-to-date, pathway-specific evaluation of epilepsy surgery outcomes in GE by combining published cases with new cases from tertiary centers. We hypothesized that surgical success rates, lesion and histopathological findings would differ by genetic pathway and sought to quantify these differences to inform presurgical epilepsy workup and counseling. Our study supports lesion-directed, genetically informed surgical candidacy in disorders with resectable substrates, quantifies the more modest yield in categories without inherent lesional phenotypes, and offers quantitative data to guide presurgical counseling, invasive evaluation, and expectation-setting.

## Methods

### Literature search

We conducted a systematic review in accordance with PRISMA guidelines. The PRISMA flowchart is included in Figure 1A. On October 25, 2025, we searched MEDLINE (via PubMed) and Scopus using the query: (epilepsy[Title/Abstract]) AND (genetic[Title/Abstract]) AND (surgery[Title/Abstract]). We accounted for publications in English with no date restrictions. Records were de-duplicated and screened in two stages (title/abstract, then full text). Two reviewers (A.I., H.K.) independently screened all abstracts and full texts.

**Figure 1.**
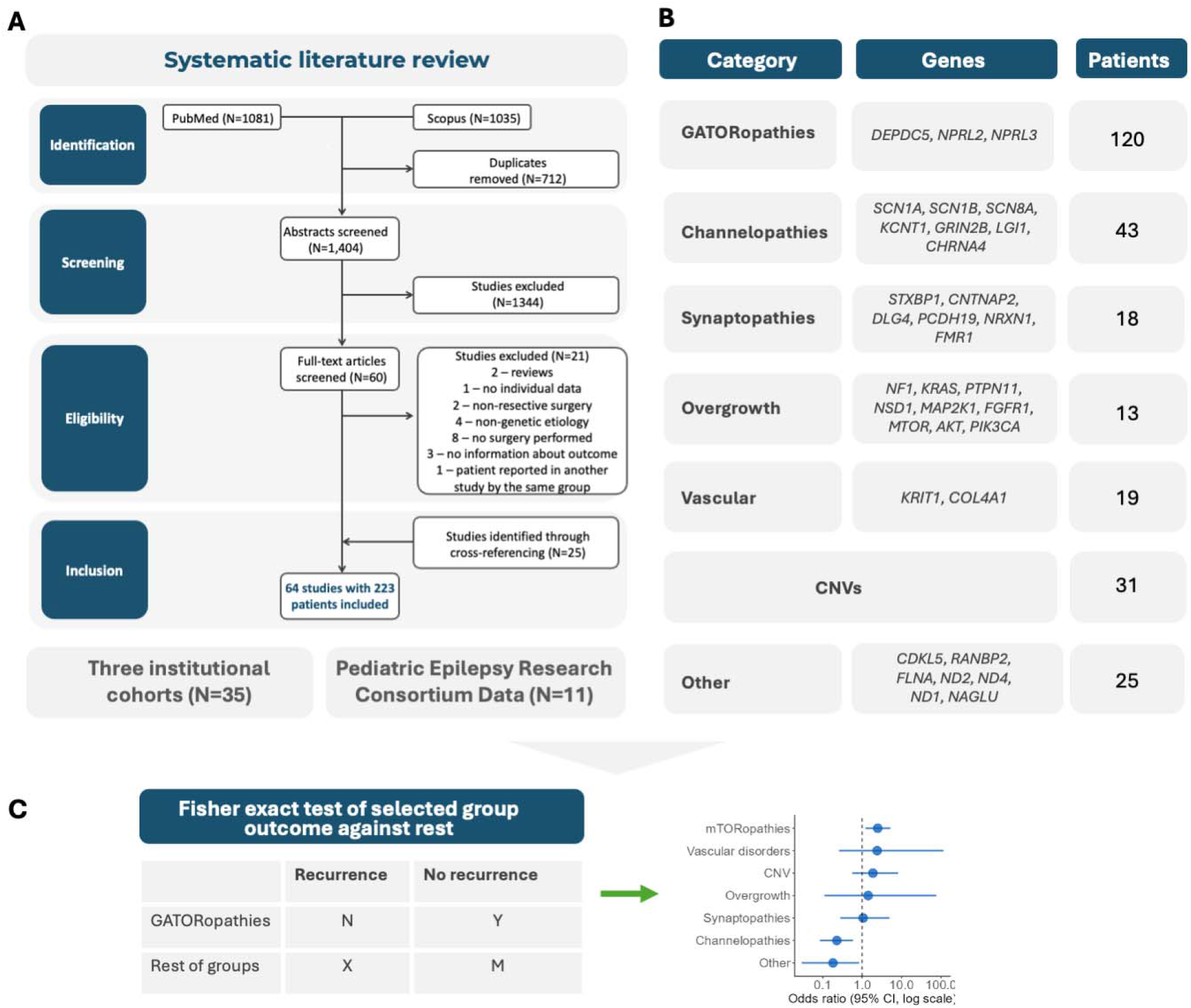
Study design overview. **A. Systematic review flowchart**. We included 64 studies contributing individual data of 223 patients. We augmented this data set with data from 35 individuals with germline genetic epilepsies who underwent surgeries within our institutions. **B. Included cases by genetic category**. We separated 6 pathway-informed groups and grouped disorders that did not fit any category into a separate group. **C. Univariate analysis outline**. We used the Fisher exact test to determine the odds of the outcome (seizure freedom and favorable outcome) in a particular genetic category compared to the rest of the genetic disorders.

Inclusion criteria were: (i) observational/interventional studies, case–control studies, case series, or case reports; (ii) participants with germline pathogenic/likely pathogenic variants in genes associated with epilepsy or copy-number variants known to be associated with epilepsy; (iii) sufficient genetic information reported to verify the variant; (iv) intervention: resective surgery or laser ablation; and (v) outcomes reported as Engel or ILAE class, seizure freedom (yes/no), or postoperative seizure frequency permitting assignment of an Engel class.

Exclusion criteria were: (i) studies reporting somatic variants only; (ii) studies in which the primary intervention was neuromodulation or callosotomy; and (iii) studies focused exclusively on tuberous sclerosis complex (TSC) or neurofibromatosis (NF), given their distinct evidence base and the risk of inflating overall outcome estimates.

The full list of included studies is available in Table S1.

### Bias assessment

Because the evidence base comprised heterogeneous observational designs, many being uncontrolled case series or single case reports, standard study-level tools (e.g., ROBINS-I, JBI/NIH checklists) were not applicable. Instead, we mitigated bias by prespecifying study eligibility criteria, summarizing missingness, and conducting sensitivity analyses, as indicated below.

### Identification of in-house cohorts

Cases of GE that underwent surgery were identified at three centers, Cleveland Clinic, Mayo Clinic, and University of Texas Health Center in Houston (UTH) using electronic health record (EHR) queries for entries containing names of genes associated with epilepsy (defined by Macnee et al. ^5^) and/or coded with International Classification of Diseases – 10^th^ Revision codes Q90-99 (“Chromosomal abnormalities, not elsewhere classified”). Cases of TSC and NF were excluded as outlined above. To identify individuals who underwent surgery among the retrieved preliminary cohorts, cases from Cleveland Clinic were matched with an institutional surgical database, and cases from Mayo Clinic were filtered for procedural codes for resective brain surgery. Cases from UTH were identified via an internal genetic case database matched with surgical records. The earliest extraction time was not limited; cases from Cleveland Clinic were extracted up to June 24, 2024, and cases from Mayo Clinic and UTH were extracted up to November 30, 2025. After retrieving individuals with potentially genetic epilepsy who underwent epilepsy surgery, their medical records were reviewed to verify the genetic diagnosis. Molecular diagnoses were verified using the current American College of Medical Genetics and Genomics/Association for Molecular Pathology interpretation guidelines for sequence variants^29^ or copy number variants.^30^

### Pediatric Epilepsy Research Consortium (PERC) data

The Pediatric Epilepsy Research Consortium (PERC) Surgery and Genetics databases are multicenter, IRB-approved registries of individuals ≤18 years (i) undergoing epilepsy surgery evaluation regardless of whether surgery is ultimately performed and (ii) with genetically determined epilepsies, respectively. The Surgery registry includes operative details and etiologies (including genetic); its analysis of genetic cases has been published in aggregate (Coryell et al., 2023)^4^ and therefore could not be included in our individual-participant review. The Genetics registry captures molecular diagnoses and treatments, but does not systematically record epilepsy surgery outcomes. The Surgery database was queried for cases meeting our inclusion criteria, and contributing sites for the Genetics registry were contacted to provide de-identified, individual-level data for patients with available surgical outcomes. We requested at minimum the variables listed in the “Extracted variables” section and accepted site-level confirmation of pathogenic/likely pathogenic germline status per local clinical standards. Mayo Clinic is a PERC-contributing institution; no cases contributed by Mayo Clinic to PERC databases were eligible for inclusion.

### Research approval

The study was approved by each individual institution’s review boards (Mayo Clinic – ID 24-007146, Cleveland Clinic – IDs 23-251 and 17-1015, and UTH – ID HSC-MS-23-1129). The IRB of reference for the PERC Genetics Database is hosted by Oregon Health & Science University and is endorsed by local IRBs of participating institutions; Surgery SIG Database data is collected under IRBs of individual participating institutions (see Table S8 for the list of institutions and Coryell et al., 2023^4^ for database specifics).

### Extracted variables

The following variables were intended to be extracted for both literature and in-house cohorts:

- Demographic (sex, age at genetic diagnosis).
- Genetic (variant information, age at genetic diagnosis).
- Surgery-related (age at intervention and type, side, and anatomical extent of intervention).
- Outcome-related (length of follow-up and either binary report of seizure freedom (yes/no), Engel score, or any seizure frequency metric that would allow assessing Engel score at last follow-up); in case of multiple surgeries, we accounted for the first surgery performed.
- Clinical (age of seizure onset, presence of MRI lesion (if present, the cases are denoted as lesional further in this paper), and pathology).

Acknowledging heterogeneity of clinical information contained in publications, we included individuals with at minimum the genetic variant, type of intervention, and Engel score/seizure frequency reported.

Genetic etiologies were grouped a priori into six biologically informed categories based on pathway annotation (PANTHER)^13^ and literature review (Figure 1B; Supplemental Methods). The disorder categories included GATORopathies (genes regulating mTOR via GATOR1, e.g. *DEPDC5, NPRL2, NPRL3*), channelopathies (ion channel genes, e.g. *SCN1A, KCNT1*), synaptopathies (synaptic function genes, e.g. *STXBP1, PCDH19*), overgrowth disorders (developmental overgrowth syndromes, e.g. PTEN hamartoma syndrome), vascular disorders (cerebrovascular developmental disorders, e.g. *COL4A1*-related porencephaly), and copy-number variants (CNVs; recurrent deletions/duplications associated with epilepsy). We distinguished between GATORopathies and other overgrowth disorders (e.g. *PTEN, AKT*) due to overlapping but distinct molecular mechanisms (mTOR pathway vs. PI3K/AKT signaling pathway), phenotypes (more common extensive cerebral malformations and systemic overgrowth features in germline disorders affecting the AKT pathway), prevalence, and clinical relevance.^14–17^ Disorders not fitting into any of the groups were combined into the other disorder group. CNVs were reclassified to a pathway category when the interval clearly encompassed a driver gene (e.g., a 16p13.3 deletion including *NPRL3* was classified as a GATORopathy); otherwise, CNVs remained in the CNV group.

Surgical extent was harmonized as temporal, extratemporal, multilobar, or hemispheric. Seizure freedom was defined as Engel I at last follow-up; favorable outcome was defined as Engel I–II.

### Statistical analysis

No prospective sample-size calculation was performed; the study was conducted as an exploratory, hypothesis-generating gene-level analysis. Descriptive statistics are reported using ratios, medians, and interquartile ranges (IQR). Between-group comparisons used the Kruskal– Wallis test for continuous variables and Fisher’s exact test for categorical variables. All tests were two-sided with α=0.05.

We performed a univariate individual-case pooled analysis. For each genetic category, the proportion achieving the outcome (seizure freedom or favorable outcome) was compared with all other categories combined using Fisher’s exact test. We report odds ratios (ORs) with 95% confidence intervals, displayed as forest plots. Analyses were conducted on an available-case basis: all cases with a non-missing outcome and category assignment were included for seizure freedom; for favorable outcome, only cases with an ascertainable Engel class were analyzed.

Given substantial heterogeneity across source reports and missingness in key covariates related to surgical prognosis, we did not fit multivariable models adjusting for all predictors to avoid unstable estimates on a markedly reduced complete-case subset. We therefore prioritized transparent univariate contrasts supplemented by sensitivity analyses. Data missingness is summarized in Figure S1.

We performed prespecified sensitivity analyses by (i) stratifying by MRI lesion status (lesional vs non-lesional); (ii) excluding GATORopathies given their over-representation and inherently focal phenotype; and (iii) restricting to literature-only cases to minimize potential practice-pattern effects from institutional cohorts.

Statistical analysis and visualization were performed in R (ver. 4.4.3) using RStudio (2025.09.2, build 418).

### Data availability

The code and literature case dataset are available on GitHub repository (https://github.com/hypsarrhythmia/gen_epi_surg). The literature case dataset is also available as Table S1.

## Results

### Cohort overview

From our systematic literature review, three institutional cohorts, and data from the PERC databases, we identified 269 patients with GE who underwent resective surgery or ablation. This cohort included 223 individual cases aggregated from 64 published studies, 35 cases from three epilepsy centers, and 11 PERC cases. Etiologies were grouped into seven biologically informed categories: GATORopathies (120 patients, 44.6%), followed by channelopathies (43 patients, 16.0%), CNVs (31 patients, 11.5%), vascular disorders (19 patients, 7.0%), synaptopathies (18 patients, 6.7%), overgrowth disorders (13 patients, 4.8%) with the remaining 25 cases (9.2%) classified as miscellaneous “other” disorders. Figure 1 illustrates the PRISMA literature selection process (panel A) and the proportional breakdown of cases by genetic category (panel B). While the distributions of patients within genetic categories within literature and institutional and PERC cohorts differed, there was no difference in the number of MRI-lesional cases or outcomes (Table S3)

### Clinical and surgical characteristics of the cohort

The median seizure onset in the total cohort was 1.1 years (IQR 0.2–5.0; Figure 2A). Among 149 patients with available data, the median age at surgery was 7.5 years (IQR 2.5–16.0) (Figure 2B), with a median delay from seizure onset to surgery of 2.8 years (IQR 0.6-7.6 years). Across genetic subgroups, the time from epilepsy onset to surgery varied, with the longest delay in channelopathies (median 6.8 years (IQR 4.4.-13.2); Table 1 and Figure 2C). Among 54 cases with available data, the median age at genetic diagnosis was 11.0 years (IQR 2.7-23.5) (Figure 2D).

**Figure 2.**
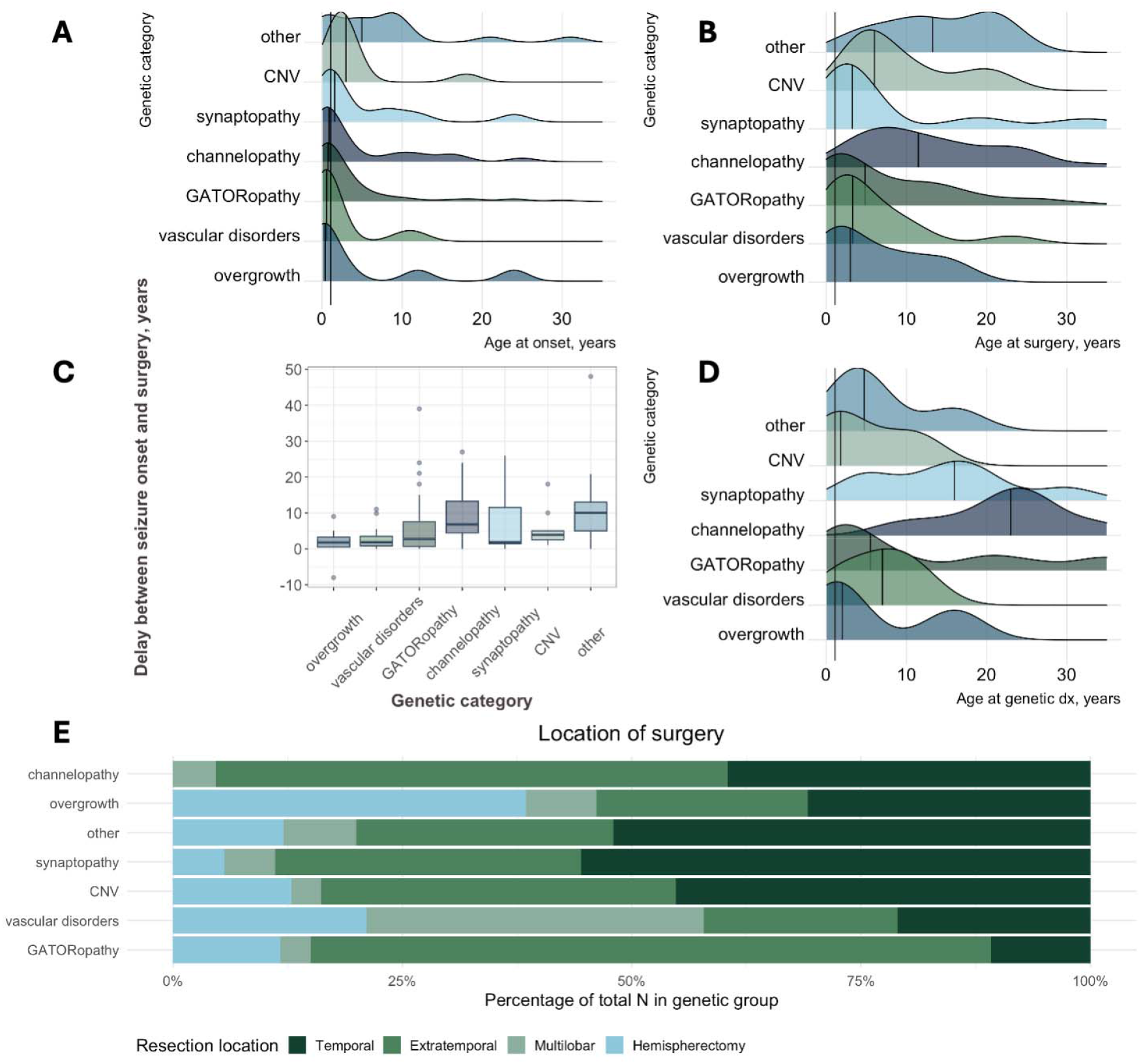
Clinical and surgical characteristics of the cohort. **A**. Ridge plot illustrating age at seizure onset. GATORopathies, vascular and overgrowth disorders, and channelopathies manifested seizures before the age of 1 year. **B**. Ridge plot showing age at surgery (available for 149 individuals). Individuals with overgrowth disorders were operated on at the youngest age. **C**. Time between seizure onset and surgery. Individuals with channelopathies had the longest delay to surgery. **D**. Age at genetic diagnosis (available for 54 individuals). Individuals with overgrowth disorders received a genetic diagnosis at the earliest in life. **E**. A bar plot showing the location of the surgery across genetic categories. Extratemporal (i.e., frontal, parietal, and occipital) surgeries prevailed in the total cohort.

**Table 1.**
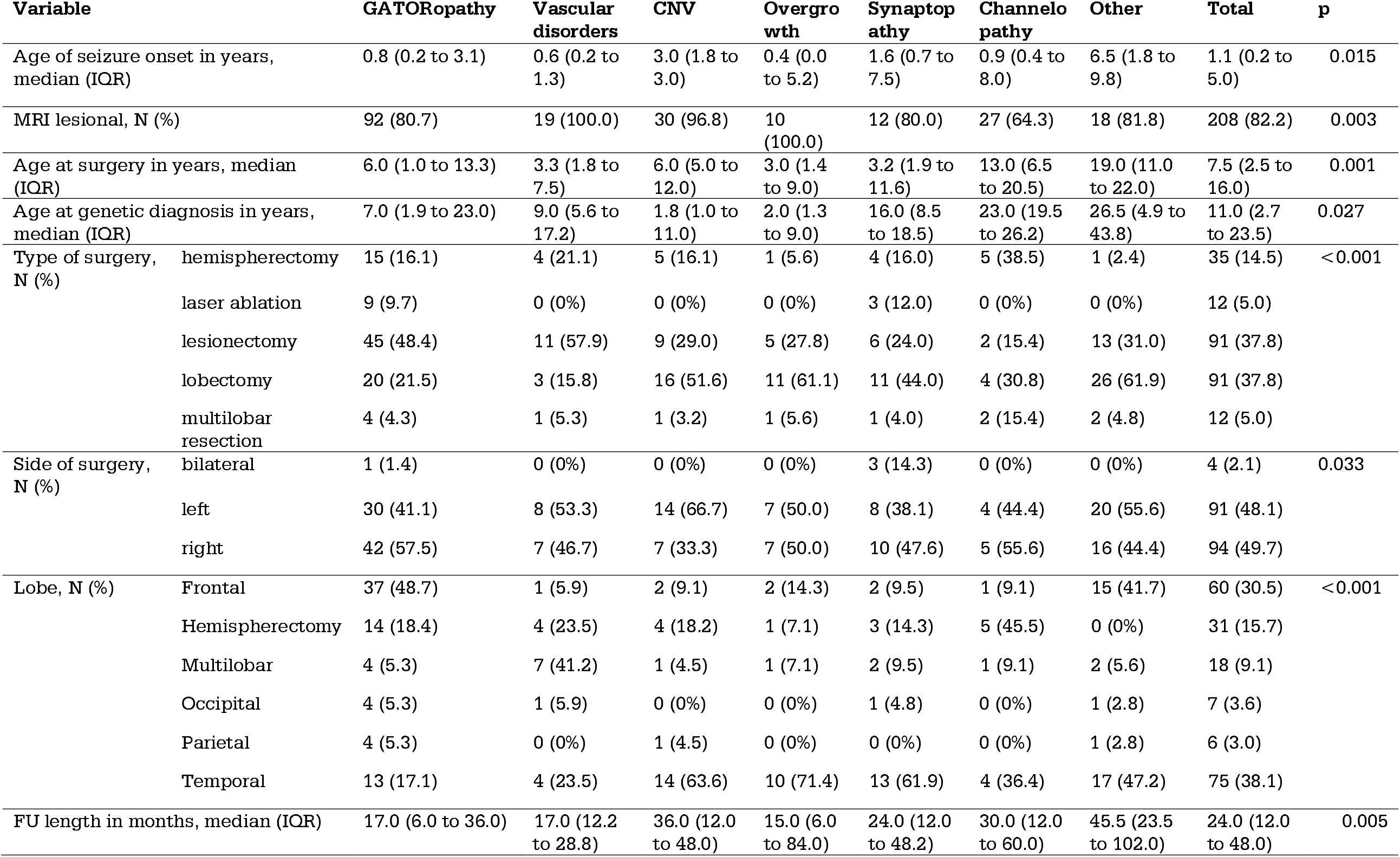
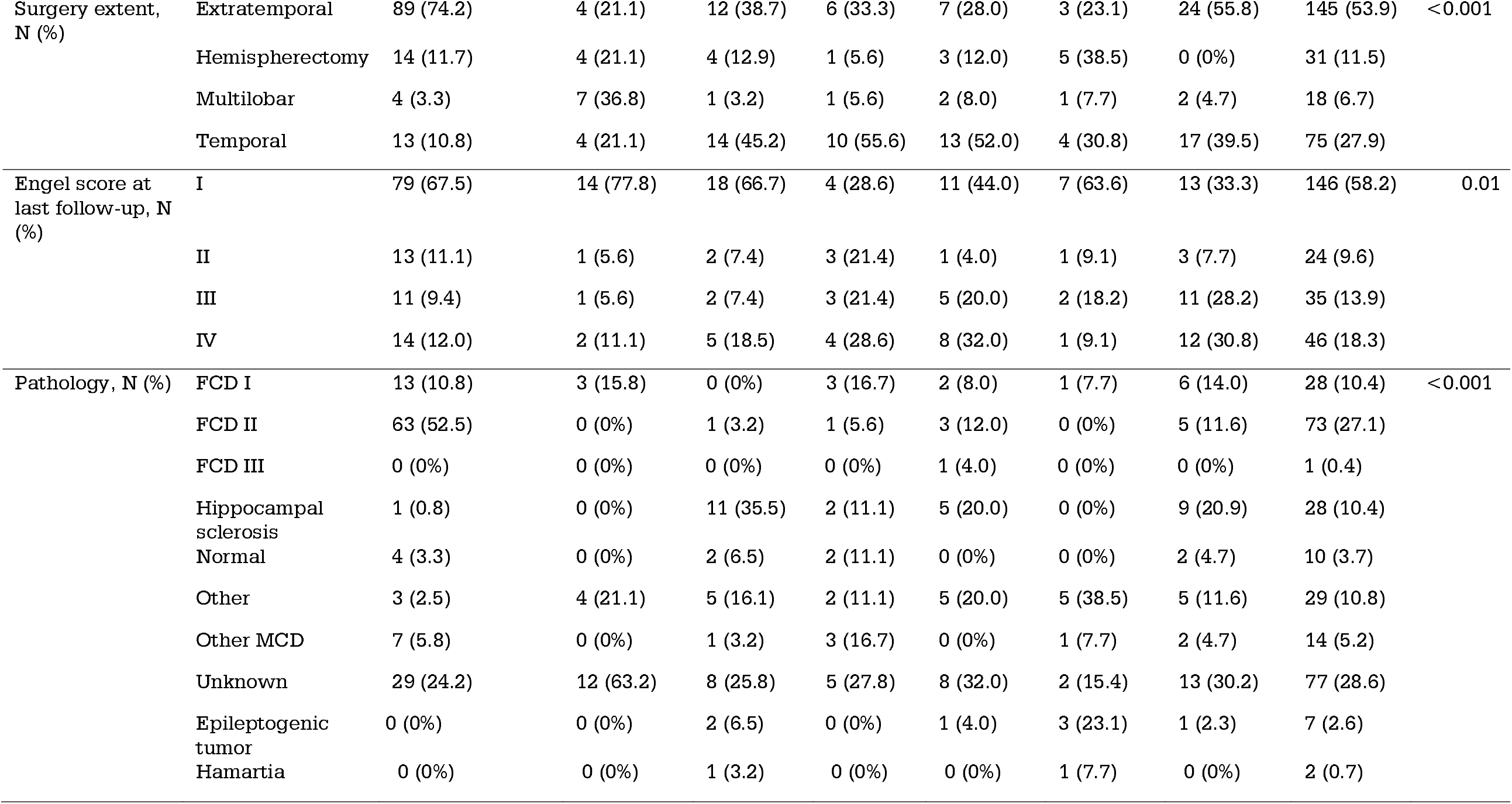
Cohort characteristics.

| Variable |  | GATORopathy | Vascular disorders | CNV | Overgrowth | Synaptopathy | Channelopathy | Other | Total | p |
| --- | --- | --- | --- | --- | --- | --- | --- | --- | --- | --- |
| Age of seizure onset in years, median (IQR) |  | 0.8 (0.2 to 3.1) | 0.6 (0.2 to 1.3) | 3.0 (1.8 to 3.0) | 0.4 (0.0 to 5.2) | 1.6 (0.7 to 7.5) | 0.9 (0.4 to 8.0) | 6.5 (1.8 to 9.8) | 1.1 (0.2 to 5.0) | 0.015 |
| MRI lesional, N (%) |  | 92 (80.7) | 19 (100.0) | 30 (96.8) | 10 (100.0) | 12 (80.0) | 27 (64.3) | 18 (81.8) | 208 (82.2) | 0.003 |
| Age at surgery in years, median (IQR) |  | 6.0 (1.0 to 13.3) | 3.3 (1.8 to 7.5) | 6.0 (5.0 to 12.0) | 3.0 (1.4 to 9.0) | 3.2 (1.9 to 11.6) | 13.0 (6.5 to 20.5) | 19.0 (11.0 to 22.0) | 7.5 (2.5 to 16.0) | 0.001 |
| Age at genetic diagnosis in years, median (IQR) |  | 7.0 (1.9 to 23.0) | 9.0 (5.6 to 17.2) | 1.8 (1.0 to 11.0) | 2.0 (1.3 to 9.0) | 16.0 (8.5 to 18.5) | 23.0 (19.5 to 26.2) | 26.5 (4.9 to 43.8) | 11.0 (2.7 to 23.5) | 0.027 |
| Type of surgery, N (%) | hemispherectomy | 15 (16.1) | 4 (21.1) | 5 (16.1) | 1 (5.6) | 4 (16.0) | 5 (38.5) | 1 (2.4) | 35 (14.5) | <0.001 |
|  | laser ablation | 9 (9.7) | 0 (0%) | 0 (0%) | 0 (0%) | 3 (12.0) | 0 (0%) | 0 (0%) | 12 (5.0) |  |
|  | lesioneectomy | 45 (48.4) | 11 (57.9) | 9 (29.0) | 5 (27.8) | 6 (24.0) | 2 (15.4) | 13 (31.0) | 91 (37.8) |  |
|  | lobectomy | 20 (21.5) | 3 (15.8) | 16 (51.6) | 11 (61.1) | 11 (44.0) | 4 (30.8) | 26 (61.9) | 91 (37.8) |  |
|  | multilobar resection | 4 (4.3) | 1 (5.3) | 1 (3.2) | 1 (5.6) | 1 (4.0) | 2 (15.4) | 2 (4.8) | 12 (5.0) |  |
| Side of surgery, N (%) | bilateral | 1 (1.4) | 0 (0%) | 0 (0%) | 0 (0%) | 3 (14.3) | 0 (0%) | 0 (0%) | 4 (2.1) | 0.033 |
|  | left | 30 (41.1) | 8 (53.3) | 14 (66.7) | 7 (50.0) | 8 (38.1) | 4 (44.4) | 20 (55.6) | 91 (48.1) |  |
|  | right | 42 (57.5) | 7 (46.7) | 7 (33.3) | 7 (50.0) | 10 (47.6) | 5 (55.6) | 16 (44.4) | 94 (49.7) |  |
| Lobe, N (%) | Frontal | 37 (48.7) | 1 (5.9) | 2 (9.1) | 2 (14.3) | 2 (9.5) | 1 (9.1) | 15 (41.7) | 60 (30.5) | <0.001 |
|  | Hemispherectomy | 14 (18.4) | 4 (23.5) | 4 (18.2) | 1 (7.1) | 3 (14.3) | 5 (45.5) | 0 (0%) | 31 (15.7) |  |
|  | Multilobar | 4 (5.3) | 7 (41.2) | 1 (4.5) | 1 (7.1) | 2 (9.5) | 1 (9.1) | 2 (5.6) | 18 (9.1) |  |
|  | Occipital | 4 (5.3) | 1 (5.9) | 0 (0%) | 0 (0%) | 1 (4.8) | 0 (0%) | 1 (2.8) | 7 (3.6) |  |
|  | Parietal | 4 (5.3) | 0 (0%) | 1 (4.5) | 0 (0%) | 0 (0%) | 0 (0%) | 1 (2.8) | 6 (3.0) |  |
|  | Temporal | 13 (17.1) | 4 (23.5) | 14 (63.6) | 10 (71.4) | 13 (61.9) | 4 (36.4) | 17 (47.2) | 75 (38.1) |  |
| FU length in months, median (IQR) |  | 17.0 (6.0 to 36.0) | 17.0 (12.2 to 28.8) | 36.0 (12.0 to 48.0) | 15.0 (6.0 to 84.0) | 24.0 (12.0 to 48.2) | 30.0 (12.0 to 60.0) | 45.5 (23.5 to 102.0) | 24.0 (12.0 to 48.0) | 0.005 |
| Surgery extent, N (%) | Extratemporal | 89 (74.2) | 4 (21.1) | 12 (38.7) | 6 (33.3) | 7 (28.0) | 3 (23.1) | 24 (55.8) | 145 (53.9) | <0.001 |
|  | Hemispherectomy | 14 (11.7) | 4 (21.1) | 4 (12.9) | 1 (5.6) | 3 (12.0) | 5 (38.5) | 0 (0%) | 31 (11.5) |  |
|  | Multilobar | 4 (3.3) | 7 (36.8) | 1 (3.2) | 1 (5.6) | 2 (8.0) | 1 (7.7) | 2 (4.7) | 18 (6.7) |  |
|  | Temporal | 13 (10.8) | 4 (21.1) | 14 (45.2) | 10 (55.6) | 13 (52.0) | 4 (30.8) | 17 (39.5) | 75 (27.9) |  |
| Engel score at last follow-up, N (%) | I | 79 (67.5) | 14 (77.8) | 18 (66.7) | 4 (28.6) | 11 (44.0) | 7 (63.6) | 13 (33.3) | 146 (58.2) | 0.01 |
|  | II | 13 (11.1) | 1 (5.6) | 2 (7.4) | 3 (21.4) | 1 (4.0) | 1 (9.1) | 3 (7.7) | 24 (9.6) |  |
|  | III | 11 (9.4) | 1 (5.6) | 2 (7.4) | 3 (21.4) | 5 (20.0) | 2 (18.2) | 11 (28.2) | 35 (13.9) |  |
|  | IV | 14 (12.0) | 2 (11.1) | 5 (18.5) | 4 (28.6) | 8 (32.0) | 1 (9.1) | 12 (30.8) | 46 (18.3) |  |
| Pathology, N (%) | FCD I | 13 (10.8) | 3 (15.8) | 0 (0%) | 3 (16.7) | 2 (8.0) | 1 (7.7) | 6 (14.0) | 28 (10.4) | <0.001 |
|  | FCD II | 63 (52.5) | 0 (0%) | 1 (3.2) | 1 (5.6) | 3 (12.0) | 0 (0%) | 5 (11.6) | 73 (27.1) |  |
|  | FCD III | 0 (0%) | 0 (0%) | 0 (0%) | 0 (0%) | 1 (4.0) | 0 (0%) | 0 (0%) | 1 (0.4) |  |
|  | Hippocampal sclerosis | 1 (0.8) | 0 (0%) | 11 (35.5) | 2 (11.1) | 5 (20.0) | 0 (0%) | 9 (20.9) | 28 (10.4) |  |
|  | Normal | 4 (3.3) | 0 (0%) | 2 (6.5) | 2 (11.1) | 0 (0%) | 0 (0%) | 2 (4.7) | 10 (3.7) |  |
|  | Other | 3 (2.5) | 4 (21.1) | 5 (16.1) | 2 (11.1) | 5 (20.0) | 5 (38.5) | 5 (11.6) | 29 (10.8) |  |
|  | Other MCD | 7 (5.8) | 0 (0%) | 1 (3.2) | 3 (16.7) | 0 (0%) | 1 (7.7) | 2 (4.7) | 14 (5.2) |  |
|  | Unknown | 29 (24.2) | 12 (63.2) | 8 (25.8) | 5 (27.8) | 8 (32.0) | 2 (15.4) | 13 (30.2) | 77 (28.6) |  |
|  | Epileptogenic tumor | 0 (0%) | 0 (0%) | 2 (6.5) | 0 (0%) | 1 (4.0) | 3 (23.1) | 1 (2.3) | 7 (2.6) |  |
|  | Hamartia | 0 (0%) | 0 (0%) | 1 (3.2) | 0 (0%) | 0 (0%) | 1 (7.7) | 0 (0%) | 2 (0.7) |  |

In terms of the extent of surgery, most cases had extratemporal (N=145, 53.9%), followed by temporal (N=75, 27.9%), hemispheric (N=31, 11.5%), and multilobar (N=18, 6.7%) interventions (Table 1, Figure 3B). Approximately 20% of patients underwent resective surgery in adulthood (34/146 cases ≥18 years old), with a median adult surgery age of 24.5 years (IQR 21.0-33.8 years) and delay from onset to surgery of 13.5 years (IQR 9.0-20.9 years). No hemispherectomies or multilobar resections were performed in this age group (Table S4).

**Figure 3.**
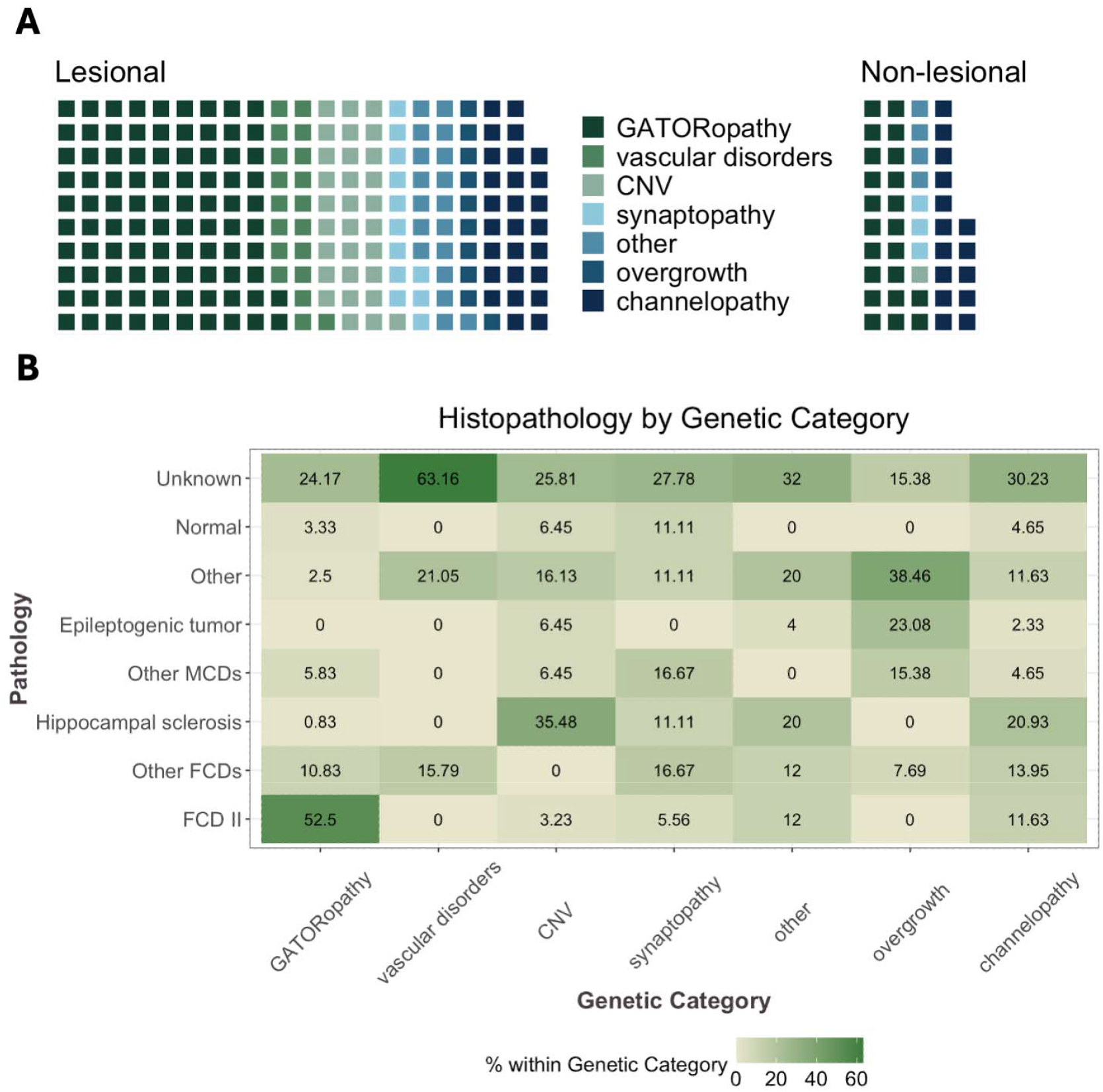
MRI and pathology characteristics. **A**. A waffle plot illustrating the number of individuals with and without lesions on MRI (information available for 253/269 individuals in the cohort). The majority of the cases were lesional across all the categories. As expected, all vascular and overgrowth disorders were lesional. **B**. Heatmap showing the distribution of reported postsurgical tissue pathologies. The number in the cells indicates the percentage of individuals in the respective genetic category. While most of the studies did not report pathology (line “Unknown”), the available reports show expected findings based on biological and clinical evidence (e.g., high prevalence of FCD type II in GATORopathies, high proportion of hippocampal sclerosis among CNVs and channelopathies that also had a high proportion of temporal surgeries).

MRI findings were available for 253 patients (94% of the cohort). Eighty-two percent of those evaluated (208/253, Table 1 and Figure 3A) had an MRI-visible structural epileptogenic lesion. As expected, categories inherently associated with focal malformations (vascular and overgrowth cases) were uniformly lesional on imaging, and 96.8% of CNV-related cases were lesional. Notably, in channelopathies and synaptopathies – disorders not classically associated with epileptogenic lesions – lesional cases accounted for 64% and 80%, respectively. Among adult surgical patients, 41.2% had no lesion on MRI, compared with only 10.7% in pediatric cases.

Histopathological data were reported for 192 individuals (71%). The distribution of histopathological diagnoses across genetic categories is shown in Table 1 and Figure 3B. The most frequent histologic diagnosis was focal cortical dysplasia type II, identified in 73 patients (∼27% of the cohort); FCD type I and hippocampal sclerosis (HS) were the next most common pathologies (28 cases (10%) each). Interestingly, HS was observed on pathology in more than 1/3 of CNV cases and in 1/5 of synaptopathies cases. Normal pathology was reported in 10 cases (4%); 6 of these were MRI-lesional.

### Surgical outcomes

At last available follow-up (median duration 24 months), 146 (58%) achieved complete seizure freedom (Engel class I) and 170 (63.0%) had a favorable outcome (Engel class I or II). Figure 4A–B illustrates the proportion of patients achieving seizure freedom and favorable outcomes in each genetic category. Outcomes differed significantly by the gene category (overall p<0.001 across groups; Table 1).

**Figure 4.**
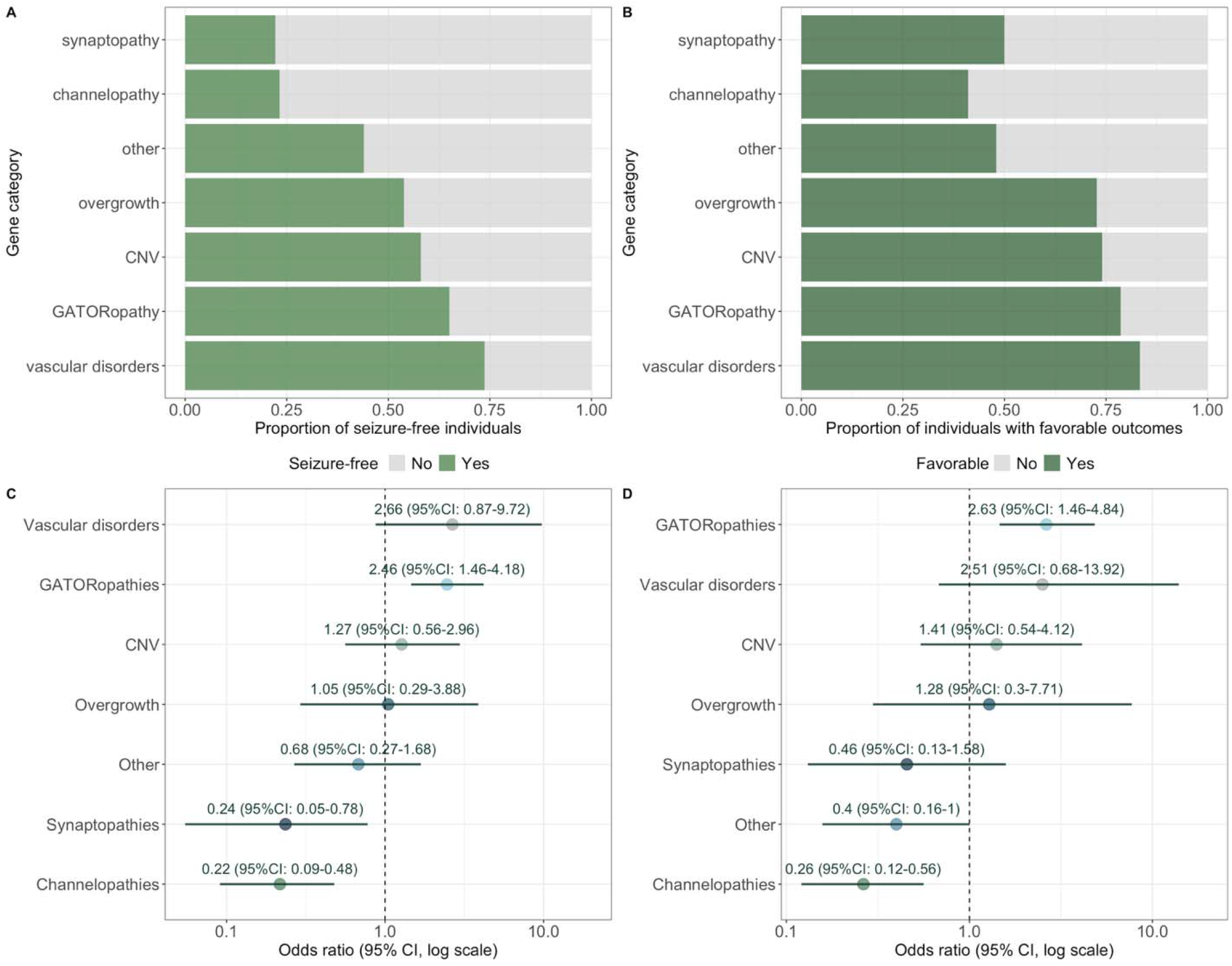
Quantitative analysis of epilepsy surgery outcomes across genetic categories. **A-B**. Bar plots illustrating the proportion of individuals achieving seizure freedom (A) and a favorable outcome (B) at the last follow-up. **C-D**. Forest plots illustrating odds of seizure freedom (C) and favorable outcome (D). The size effect is derived by comparing the outcomes in a given category against the rest of the categories taken together using Fisher’s exact test.

In univariate, category-wise comparisons of each category against all others, individuals with vascular disorders and GATORopathies were 2.66 and 2.46 times more likely to be seizure free (95% CI 0.87–9.72 and 1.46-4.18, respectively), whereas synaptopathies and channelopathies were approximately 5 times less likely to be seizure free (OR 0.24, 95% CI 0.05–0.78 and OR 0.22, 95% CI 0.09–0.48, respectively) (Figure 4C).

Results were directionally consistent across prespecified sensitivity analyses. Tables S5-7 feature comparisons between genetic categories with filtering applied for each sensitivity analysis. In lesional-only analyses, GATORopathies (OR 2.32, 95% CI 1.27–4.3) and vascular disorders (OR 2.23, 95% CI 0.72–8.24) had the highest odds (Figure S3A). Restricting to literature-only cases, odds of seizure freedom remained highest in vascular disorders (OR 3.8, 95% CI 0.99–21.55), followed by GATORopathies (OR 2.22, 95% CI 1.27–3.95) and CNVs (OR 2.01, 95% CI 0.74–6.1) (Figure S3B). Excluding GATORopathies, vascular disorders retained the highest seizure freedom odds (OR 4.43, 95% CI 1.4–16.72), whereas channelopathies - the lowest (OR 0.29, 95% CI 0.12–0.69) (Figure S2C).

Findings were similar for the favorable outcome endpoint: GATORopathies showed higher odds (OR 2.63, 95% CI 1.46–4.84), whereas channelopathies showed lower odds (OR 0.16, 95% CI 0.12–0.56) (Figure 4D), with parallel patterns in the sensitivity analyses (Figure S3A-C).

### Surgical outcomes in distinct genetic categories

#### Vascular disorders

The following genes were included in the vascular disorders category: *COL4A1* (N=9), *COL4A2* (N=6), and *KRIT1* (N=4). All cases were MRI-lesional. Procedures were predominantly lesionectomies (n=11/19, 58%), followed by hemispherectomies (n=4/19, 21%). At a median follow-up of 17 months (IQR 12.2–28.8), 14/19 (73.6%) were seizure-free; the four without clinical improvement carried variants in *COL4A1* (n=2) or *KRIT1* (n=2).

#### GATORopathies

GATORopathies were represented by *NPRL2* (N=7), *NPRL3* (N=45), *DEPDC5* (N=67). Extratemporal resections constituted 74.2% of procedures. Seizure freedom and favorable outcomes were 67.5% (79/120) and 76.7% (92/120), respectively, at a median follow-up of 17 months (IQR 6.0–36.0). There was no statistically significant difference in the proportion of seizure-free individuals on the individual gene level (*NPRL2* 5/7,71.4%; *NPRL3* 29/45, 64.4%; *DEPDC5* 44/67, 65.7%; p=1; Figure S4). Favorable outcomes were achieved in 92 (76.7%) of GATORopathies cases, of which 48 had *DEPDC5* (71.6% of all *DEPDC5* cases), 6 – *NPRL2* (85.7%), and 38 – *NPRL3* (84.4%).

#### Overgrowth disorders

Overgrowth disorders included three cases of *AKT3*, two cases of *FGFR1* and *KRAS*, and single cases of *GLI3, MAP2K1, NSD1, PTEN, PTPN11*, and *TGFBR2*. All the cases were lesional. At a median follow-up of 30 months (IQR 12.0-60.0), 7/13 (53.8%) individuals were seizure-free, and 8/13 (61.5%) had a favorable outcome.

#### Synaptopathies

This group comprised *PCDH19* (n=8), *CNTNAP2* (n=6), *STXBP1* (n=2), *DLG4* (n=1), and *NRXN1* (n=1). Among those with imaging data, 12/15 (80%) were lesional. At a median follow-up of 29 months (IQR 6–132), among those with available respective outcomes, 4/18 (22.2%) achieved seizure freedom and 7/14 (50%) had favorable outcomes. Histopathology (available in 13) ranged from FCD I/II and mMCD to hippocampal sclerosis, with 2/13 (11.1%) reported as unremarkable (Table 1).

#### Channelopathies

Channelopathies included *SCN1A* (n=25), *KCNT1* (n=3), *SCN1B* (n=2), *GRIN2B* (n=2), and single cases of *SCN2A, SCN8A, CACNA1A, HCN1, KCNA1, KCNA2, KCNH2, KCNJ10, NALCN, CHRNA4*, and *LGI1*. MRI lesions were present in 27 cases (64.2%). Pathology (available in 25) most commonly showed hippocampal sclerosis (n=9), with FCD I (n=6) and FCD II (n=5) also observed. At a median follow-up of 45 months (IQR 23.5–102), 13/43 (33.3%) were seizure-free, and 16/43 (38.1%) had favorable outcomes.

#### Copy Number Variants (CNVs)

Detailed CNVs are listed in the Supplement. MRI lesions were identified in 30/31 (96.8%); one case, a patient with ring chromosome 20 who underwent left frontal resection, lacked an MRI-visible lesion. At a median follow-up of 36 months (IQR 12.0-48.0), 18/31 (66.7%) achieved seizure freedom, and 20/31 (64.5%) had favorable outcomes.

#### Other disorders

Etiologies not attributable to the predefined categories were grouped as Other (the list of genes is included in Figure 1 and Supplemental Methods, and individual literature cases are available in Table S2). Outcomes were generally unfavorable in *CDKL5* deficiency disorder (3 cases; two temporal resections and one hemispherectomy; all Engel IV) and in mitochondrial disorders (4/5 Engel III–IV; one *ND1* case with hippocampectomy for hippocampal sclerosis achieved sustained seizure freedom at 3 years). In *RANBP2*-related acute necrotizing encephalopathy, functional hemispherectomy aborted refractory status epilepticus in one of two cases.

## Discussion

As genetic testing moves upstream in the presurgical work-up, the central question is no longer whether individuals with germline genetic epilepsies (GE) can be operated on, but in which germline disorders resection is likely to be curative rather than palliative. In this individual-participant analysis of 269 patients with germline pathogenic variants who underwent resective surgery or ablation, combining 223 published cases with 46 cases from three tertiary epilepsy centers and Pediatric Epilepsy Research Consortium databases, we provide to-date the most comprehensive, pathway-informed view of surgical outcomes in GE. Approximately half of patients achieved seizure freedom, and nearly two-thirds had a favorable outcome at a median of two years follow-up. Structural substrates were identified on MRI in over 80% of evaluable cases, and focal cortical dysplasia type II was the single most common histopathology, indicating that surgical candidacy for individuals with GE was likely driven by the presence of an operable lesion on MRI. By organizing etiologies into biologically coherent pathways, we demonstrate that disorders with inherently focal, resectable substrates-GATORopathies, vascular malformations, overgrowth syndromes and many copy-number variants-show Engel I rates in the range of ∼55–75% whereas channelopathies and synaptopathies, despite the presence of MRI-identifiable lesions in over 60% and 80% of cases respectively, rarely exceed 20–30% seizure freedom and have significantly lower odds of seizure freedom and favorable outcome than other categories. These pathway-specific estimates extend and refine earlier, smaller series and mixed somatic–germline meta-analyses.^5,12^ They remain stable across sensitivity analyses restricted to lesional cases, to literature-only cases, and excluding GATORopathies. Our findings suggest that genetic testing should be integrated early as a stratifying variable rather than a contraindication in drug-resistant epilepsy workup and counseling.

Our study updates and extends prior syntheses of surgical outcomes in genetic epilepsies.^5,12^ We restricted the analysis to germline etiologies because clinical genetic testing for germline variants is widely available and feasible during the presurgical workup, making results directly applicable to contemporary practice.^18^ In contrast, detection of somatic variants typically requires resected tissue or research-grade assays with limited preoperative availability, and would introduce mechanistic heterogeneity that could obscure pathway-specific inferences. Despite this narrower scope—and an overall cohort that was predominantly MRI-lesional—the observed seizure-freedom rate was comparable to earlier estimates (approximately 52%–56%). Whereas one prior meta-analysis did not stratify outcomes by genetic pathway^12^, our pathway-based approach (and the earlier review that did)^5^ demonstrates consistent differences: higher odds of seizure freedom in GATOR/mTOR-related and vascular disorders, and lower odds in channelopathies and synaptopathies. Variations between studies likely reflect differences in case mix (age, lesion status), surgical era and extent, and follow-up duration, but the direction of the pathway-specific signal appears stable across analyses.

Our cohort was enriched for GATORopathies compared to other genetic etiologies. There are several factors plausibly driving this over-representation. First, germline variants in GATOR-complex genes (e.g., *DEPDC5, NPRL2/3*) are associated with mTOR-pathway dysregulation and a spectrum of malformations of cortical development; the resulting lesional phenotype aligns with conventional surgical candidacy and yields higher selection into operative cohorts.^2,19^ Second, also applicable to vascular disorders in our dataset (e.g., *COL4A1/2, KRIT1*), many GATORopathy variants are not de novo but rather inherited in an autosomal dominant mode, often with incomplete penetrance.^16,17,20^ This facilitates familial transmission, enlarging the pool of affected individuals presenting for surgical evaluation. In contrast, most synaptopathy and channelopathy variants are de novo, with parental variant inheritance present in the utmost minority of cases.^21–23^

Categories characterized by discrete, resectable substrates (GATORopathies and vascular disorders) achieved the highest rates of seizure freedom and favorable outcomes. Notably, the categories with the best outcomes (vascular disorders and CNVs) more often underwent extensive procedures (multilobar or hemispheric resections). This pattern is biologically plausible—broader resections may encompass a larger epileptogenic anatomical substrate or network—but also raises the possibility of confounding by indication, whereby the presence of more extensive lesions, such as porencephaly in *COL4A1*-related disorders, prompts more aggressive surgery and, in turn, higher success rates.^24^

Notably, seizure freedom in GATORopathies is not universal. Our estimate exceeds that of prior meta-analysis that separated GATORopathies (germline only) as a group^5^, yet falls below rates reported in a pediatric, single-country series.^25^ These differences likely reflect case mix, age, selection criteria, and follow-up duration across studies. Variability in outcomes is plausibly influenced by technical factors (lesion detectability, extent and margins of resection, and use of invasive monitoring) and by biology that extends beyond the MRI-visible lesion. Experimental models of *DEPDC5*-related mTOR activation indicate lowered seizure threshold and molecular hallmarks of focal cortical dysplasia even in cortex with preserved lamination^26^, raising the possibility of MRI-negative or perilesional epileptogenic tissue that limits complete resection.

Most cases in this analysis were lesional—even within channelopathies and synaptopathies, which are not classically defined by structural lesions. FCD type I has been described in association with variants such as *SCN1A, KCNT1, PCDH19*;^27–30^ its subtle imaging features and lack of characteristic EEG patterns make presurgical localization challenging and are linked to less favorable postoperative outcomes.^31^ Hippocampal sclerosis may represent a consequence of long-standing seizures rather than the primary genetic substrate in these categories of genetic disorders. It is a plausible hypothesis that removing an additional epileptogenic lesion, even given it is not directly related to the pathogenic effect of causative genetic variant, could yield palliative effect and relieve burden of disease; yet, approximately half of surgically treated individuals in these categories had Engel III–IV outcomes in our compilation. Together, these observations indirectly support the view that channelopathies and synaptopathies reflect distributed network hyperexcitability; a focal resection may therefore be palliative rather than curative. Notably, within the “Other” category, which included disorders without an inherent lesional component (e.g., *CDKL5*, mitochondrial, *RANBP2*), outcomes were generally unfavorable, reinforcing that diffuse genetic developmental encephalopathies seldom yield durable seizure freedom. Alternative strategies—such as neuromodulation—warrant consideration and further study in this population.^32^

The majority of cases in our study were pediatric – an expected finding given that most genetic epilepsies present in childhood and current clinical genetic testing practices primarily consider pediatric patients. Adult genetic epilepsy surgery candidates were rarer and tended to lack MRI lesions, which could contribute to worse outcomes in that subgroup. This underscores the importance of early intervention: indeed, our data hint that shorter epilepsy duration before surgery (as seen in GATORopathies and overgrowth disorders) may partly explain their better outcomes, echoing general epilepsy surgery literature that earlier surgery yields higher success rates.^2^

Only a minority of included cases reported the timing of genetic testing. Nonetheless, genetic testing is increasingly integrated into presurgical evaluation and tends to redirect management away from resection in case of identification of disorders characterized by inherently diffuse epileptogenicity, such as channelopathies and synaptopathies, even in the presence of focality.^4,9,11,19^ Our data add quantitative context to this practice: pathway-specific differences in surgical yield suggest that a molecular diagnosis can refine pretest probability of seizure freedom, calibrate the threshold for invasive monitoring, and inform counseling about curative versus palliative intent.

This study has several limitations inherent to literature-based individual-participant synthesis. Publication and selection bias are likely: successful or MRI-lesional cases are more likely to be reported. In particular, the unexpectedly high proportion of lesional cases in channelopathy/synaptopathy likely reflects selection for surgically ‘promising’ phenotypes, limiting generalizability to the broader, largely non-lesional population. In addition, earlier genetic testing practice included single-gene sequencing, and thus disorders that were specifically tested for are more likely to be represented. We could not ascertain the denominator of genetically diagnosed patients evaluated but not operated on. Heterogeneity exists in presurgical evaluation; clinical variables were inconsistently reported, leading to variable missingness and restricting the scope of analyses. Because missingness was nontrivial, we prioritized transparent univariate comparisons with prespecified sensitivity analyses and avoided over-interpreting multivariable models on a much smaller complete-case subset. As such, residual confounding by unmeasured or incompletely measured factors (e.g., dual genetic etiologies or somatic variants, seizure semiology, medication history, comorbidities) is possible. Small numbers within several genetic strata produced wide confidence intervals and limited inference at the gene level. To preserve power, we grouped etiologies into pathway-based categories and retained an “other” class; while pragmatic, this aggregation risks obscuring gene-specific effects. Misclassification is also possible for CNVs that encompass multiple candidate genes or when variant interpretation evolved over time.

Despite these limitations, this study provides the largest germline-focused, pathway-aware aggregation to date, applies prespecified sensitivity analyses that reproduce the core signal, and offers clinically actionable stratification of surgical yield across genetic pathways to guide presurgical counseling and future prospective studies.

## Conclusions

Our systematic review of outcomes in individuals with GE undergoing epilepsy surgery reinforces previous observations of mostly favorable outcomes in GATORopathies and other disorders characterized by the presence of a clear epileptogenic lesion, such as vascular disorders and overgrowth disorders. In addition, it highlights disorders characterized by diffuse epileptogenicity, such as channelopathies and synaptopathies, as less favorable compared to other surgeries; in these categories, even resection of a co-occurring MRI lesion (hippocampal sclerosis, FCD) did not lead to a favorable outcome in majority of cases. Genetic testing can be a valuable tool in pre-surgical epilepsy assessment as it may shorten the time to the appropriate pathway and prevent non-beneficial resections. Prospective, genotype-aware surgical registries with standardized reporting are required to inform precision interventions in GE.

## Funding

The study did not receive external funding.

## Conflicts of Interest

The authors report no conflicts of interest relevant to this work.

## Author contributions

A.I. conceived and designed the study, collected data, performed the analyses and visualizations, drafted the initial manuscript, and led manuscript revisions. S.B., H.Y.K., J.C., J.S., and S.P. contributed to data collection and participated in drafting and revising the manuscript. A.F. provided supervision and contributed to drafting and revising the manuscript. The PERC Surgery Workgroup and PERC Genetics Workgroup contributed data collection. D.L. contributed to study design, supervision, data curation, and drafting and revising the manuscript. All other authors contributed to drafting and revising the manuscript and approved the final version.

## Notes

### Competing Interest Statement

Dennis Lal, PhD received institutional research funding from the National Institutes of Health (NIH), National Institute of Neurological Disorders and Stroke (NINDS) (R01 NS117544).
Elia Pestana-Knight, MD has received consulting and speaker fees from Marinus Pharmaceuticals, Pyrus Pharmaceuticals, Takeda Pharmaceuticals, BioMarin Pharmaceuticals, UCB, and Tenecia Biotech.
Imad Najm, MD serves on the advisory board and speakers bureau for SK Life.
Elaine Wirrell, MD serves on the Data and Safety Monitoring Board (DSMB) for Encoded and GRIN Pharma and has received consulting fees from Biocodex and Jazz Pharmaceuticals.
All other authors report no disclosures.

### Funding Statement

This study did not receive any funding.

### Author Declarations

The study was approved by each individual institution's review boards (Mayo Clinic - ID 24-007146, Cleveland Clinic - IDs 23-251 and 17-1015, and The University of Texas Health Science Center at Houston - ID HSC-MS-23-1129). The IRB of reference for the PERC Genetics Database is hosted by Oregon Health & Science University and is endorsed by local IRBs of participating institutions; Surgery SIG Database data is collected under IRBs of individual participating institutions.

