## Supplemental materials for "Germline genetic variants and epilepsy surgery response: individual-participant pooled analysis of 269 patients"

Alina Ivaniuk, MD^1^; Sunanjay Bajaj^2^, Christian M Boßelmann, MD^3^; Hyun Yong Koh MD, PhD^4^; Elia Pestana-Knight, MD^5^; Xiaoming Zhang, PhD^5^; William Bingaman, MD^5,6^; Imad Najm, MD^5^; Manish Shah, MD^7^; Nitin Tandon^7^, MD; Gretchen Von Allmen^8^, MD; Samden D. Lhatoo, MD^2^; William Tatum, DO^1^; Brin Freund, MD^1^; Kai J. Miller, MD, PhD^9^; Elaine Wirrell, MD^10^; Anthony Fine, MD^10^; Jason Coryell^11^; John Schreiber^12^; Scott Perry^13^; Pediatric Epilepsy Research Consortium (PERC) Surgery Workgroup; Pediatric Epilepsy Research Consortium (PERC) Genetics Workgroup; Dennis Lal, PhD^13,14^

^1^Department of Neurology, Mayo Clinic Florida, Jacksonville, FL, USA

^2^Department of Neurology, McGovern Medical School, The University of Texas Health Science Center at Houston, Houston, TX, USA

^3^Department of Neurology and Epileptology, University Hospital Tübingen, Tübingen, Germany

^4^Department of Pediatrics, Texas Children's Hospital, Houston, TX, USA

^5^Epilepsy Center, Neurological Institute, Cleveland Clinic, Cleveland, OH, USA

^6^Department of Neurosurgery, Neurological Institute, Cleveland Clinic, Cleveland, OH, USA

^7^Department of Neurosurgery, McGovern Medical School, The University of Texas Health Science Center at Houston, Houston, TX, USA

^8^Department of Pediatrics, McGovern Medical School, The University of Texas Health Science Center at Houston, Houston, TX, USA

^9^Department of Neurologic Surgery, Mayo Clinic, Rochester, MN, USA

^10^Department of Neurology, Division of Epilepsy, Mayo Clinic, Rochester, MN, USA

^11^Department of Pediatrics, Oregon Health and Sciences University, Portland, OR, USA

^12^Department of Epilepsy, Neurophysiology, and Critical Care Neurology, Children's National Hospital, Washington, DC, USA

^13^ Department of Neurology, Jane and John Justin Institute for Mind Health, Cook Children's Medical Center, Fort Worth, TX, USA

^14^ Center for Innovation in Health Informatics, The University of Texas at Arlington, Arlington, TX, US

### Supplemental Methods

We reviewed literature on biological function and clinical manifestations of each recorded gene and assigned them to the following categories: GATORopathies, channelopathies, synaptic formation disorders, synaptic transmission disorders, overgrowth disorders, vascular disorders, and other disorders. The following category assignment was used:

| Category | Genes | Sources |
| --- | --- | --- |
| GATORopathies | *DEPDC5, NPRL2, NPRL3* | [Auvin and Baulac, 2022](https://www.sciencedirect.com/science/article/pii/S0035378723008676); [Nguen and Bordey, 2021](https://www-frontiersin-org.ccmain.ohionet.org/articles/10.3389/fnana.2021.664695/full); [Baldassari et al., 2019](https://www.gimjournal.org/article/S1098-3600(21)04626-8/fulltext), |
| Channelopathies | *SCN1A, SCN1B, SCN8A, KCNT1, GRIN2B, LGI1, CHRNA4* | [Brunklaus et al., 2022](https://academic.oup.com/brain/article/145/12/4275/6509259); [Musto, Gardella, Møller, 2019](https://www.ejpn-journal.com/article/S1090-3798(19)30429-5/abstract); [Johannesen et al., 2021](https://academic.oup.com/brain/article/145/9/2991/6357698); [Bonardi et al., 2021](https://academic.oup.com/brain/article/144/12/3635/6296590); [Platzer et al., 2017](https://jmg.bmj.com/content/54/7/460); [Lerche et al., 2013](https://physoc.onlinelibrary.wiley.com/doi/10.1113/jphysiol.2012.240606); [Gooley, Crompton, and Berkovic 2022](https://onlinelibrary.wiley.com/doi/10.1684/epd.2021.1393) |
| Synaptopathies | *STXBP1, CNTNAP2, DLG4, PCDH19, NRXN1, FMR1* | [Verhage and Sørensen, 2020](https://www.cell.com/neuron/fulltext/S0896-6273(20)30405-0); [Lazaro et al., 2019](https://www.cell.com/cell-reports/fulltext/S2211-1247(19)30617-5); [Giansante et al., 2021](https://www.nature.com/articles/s41380-023-02022-1); [Kwong et al., 2015](https://journals.plos.org/plosone/article?id=10.1371/journal.pone.0126446); [Varea et al., 2015](https://www.ncbi.nlm.nih.gov/pmc/articles/PMC4434727/); [Pfleiffer and Huber, 2007](https://www.jneurosci.org/content/27/12/3120) |
| Overgrowth disorders | *NF1, KRAS, PTPN11, NSD1, MAP2K1, FGFR1, MTOR, AKT, PIK3CA, PTEN* | [Corral et al., 2003](https://onlinelibrary.wiley.com/doi/10.1002/jcp.10349); [Diociaiuti et al., 2022](https://www.mdpi.com/2227-9059/10/6/1460); [Kim et al., 2022](https://bmcmedgenomics.biomedcentral.com/articles/10.1186/s12920-022-01362-1); [Huang et al., 2014](https://www.eurekaselect.com/article/61461); [Tartaglia et al., 2006](https://www.cell.com/ajhg/fulltext/S0002-9297(07)62359-3); [Sun et al., 2023](https://www.cell.com/molecular-cell/fulltext/S1097-2765(23)00430-6); [Shen et al., 2019](https://jmg.bmj.com/content/57/11/794); [Pierpont, Semrud-Clikeman, and Pierpont, 2019](https://onlinelibrary.wiley.com/doi/10.1002/ajmg.a.38044); [Paradiso, Zucchini, and Simonato, 2013](https://www.ncbi.nlm.nih.gov/pmc/articles/PMC3772316/); [Bennett et al., 2016](https://www.sciencedirect.com/science/article/pii/S0002929716000574), [Moloney, Cavalleri, and Delanty, 2021](https://academic.oup.com/braincomms/article/3/4/fcab222/6375444); [Dobyns and Mirzaa, 2019](https://onlinelibrary.wiley.com/doi/10.1002/ajmg.c.31736) |
| Vascular disorders | *KRIT1, COL4A1, COL4A2* | [Draheim et al., 2014](https://journals.biologists.com/jcs/article/127/4/701/54828/Cerebral-cavernous-malformation-proteins-at-a); [Swamy and Glading, 202](https://www.frontiersin.org/articles/10.3389/fcvm.2022.954780/full)2; [Lanfrankoni and Markus, 2010](https://www.ahajournals.org/doi/10.1161/STROKEAHA.110.581918); [Bersano et al., 2021](https://link.springer.com/article/10.1007/s00415-020-09836-x); [Meuwissen et al., 2015](https://linkinghub.elsevier.com/retrieve/pii/S1098360021031476) |
| Copy number variants (CNVs) | All copy number variants | N/A |
| Other | *CDKL5* (kinase), *RANBP2* (GTP-binding protein), *FLNA* (neuronal migration), *ND2, ND4, ND1* (mitochondrial), *NAGLU* (mucopolysaccharidosis) | [Zhu et al., 2023](https://www.cell.com/cell-reports/fulltext/S2211-1247(23)01214-7); [Jagtap et al., 2019](https://academic.oup.com/hmg/article/28/21/3625/5569474); [Moreno-Oñate et al, 2020](https://www.sciencedirect.com/science/article/pii/S0022283620302539?via%3Dihub); [Jiang et al., 2022](https://www.mdpi.com/1422-0067/23/7/3548); [Zhang et al., 2013](https://www.jneurosci.org/content/33/40/15735); [Vriend and Oegma, 2021](https://www.ejpn-journal.com/article/S1090-3798(21)00182-3/fulltext); [Vartak et al., 2015](https://www.sciencedirect.com/science/article/pii/S0925443915001143); [Lim et al., 201](https://faseb.onlinelibrary.wiley.com/doi/epdf/10.1096/fj.201500137R)6; [Li et al., 2020](https://www.sciencedirect.com/science/article/pii/S1043661820313888?via%3Dihub); [Yogalingam and Hopwood, 2001](https://onlinelibrary.wiley.com/doi/10.1002/humu.1189)[; El Fatimy et al., 2022](https://molecular-cancer.biomedcentral.com/articles/10.1186/s12943-022-01494-z) |

### Supplemental Figures

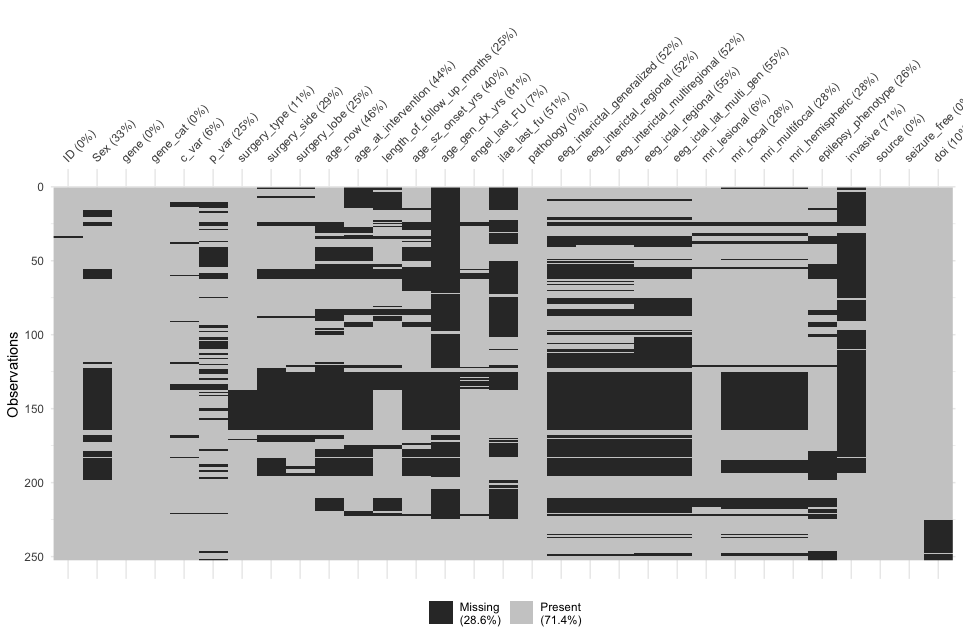

Supplemental Figure 1. Patterns of missing data in the collected data set. The Y axis represents each included case, and the X axis lists data set variables. Darker hue of gray indicates a missing variable.

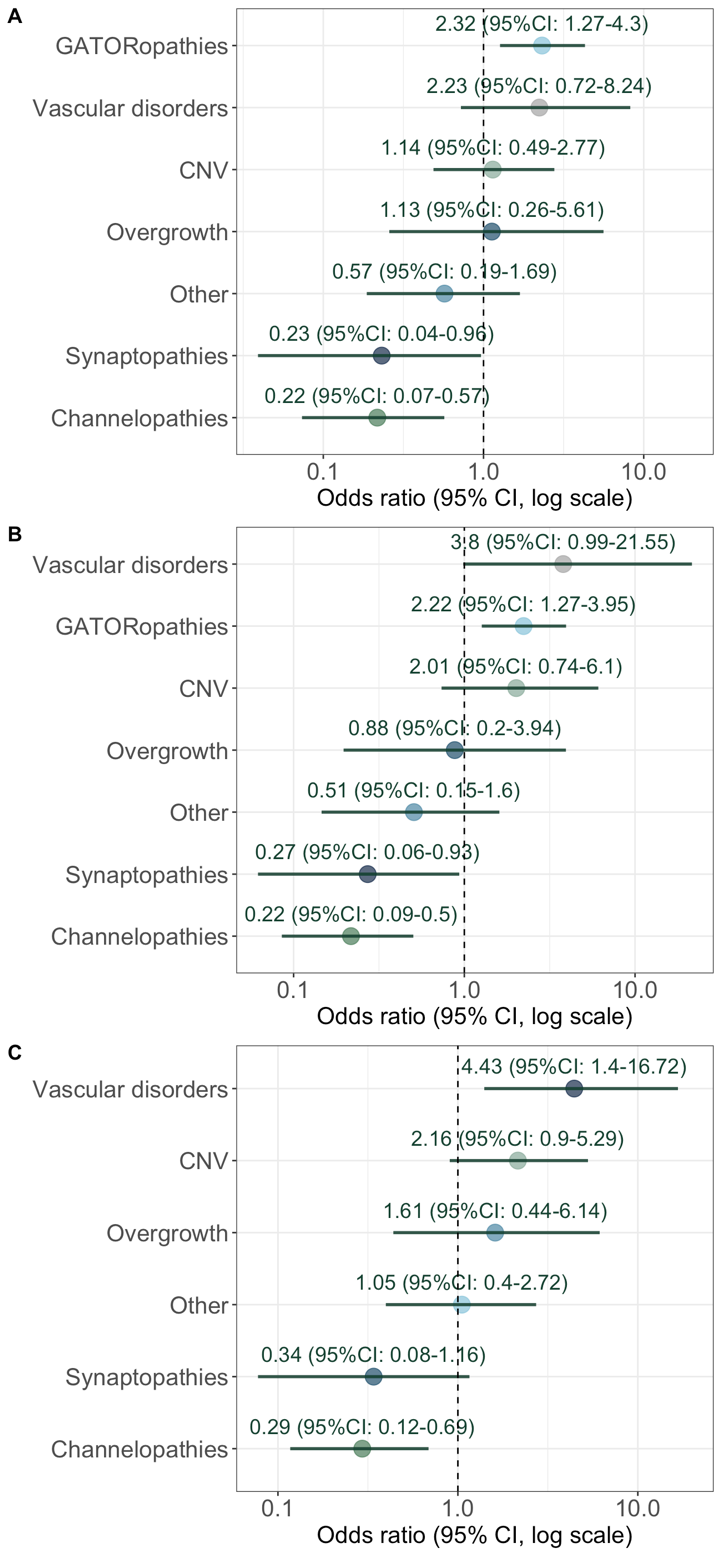

Supplemental Figure 2. Univariate analysis of association of genetic categories with seizure freedom in subcohorts: A) lesional cases only; B) literature cases only; C) all cases besides GATORopathies.

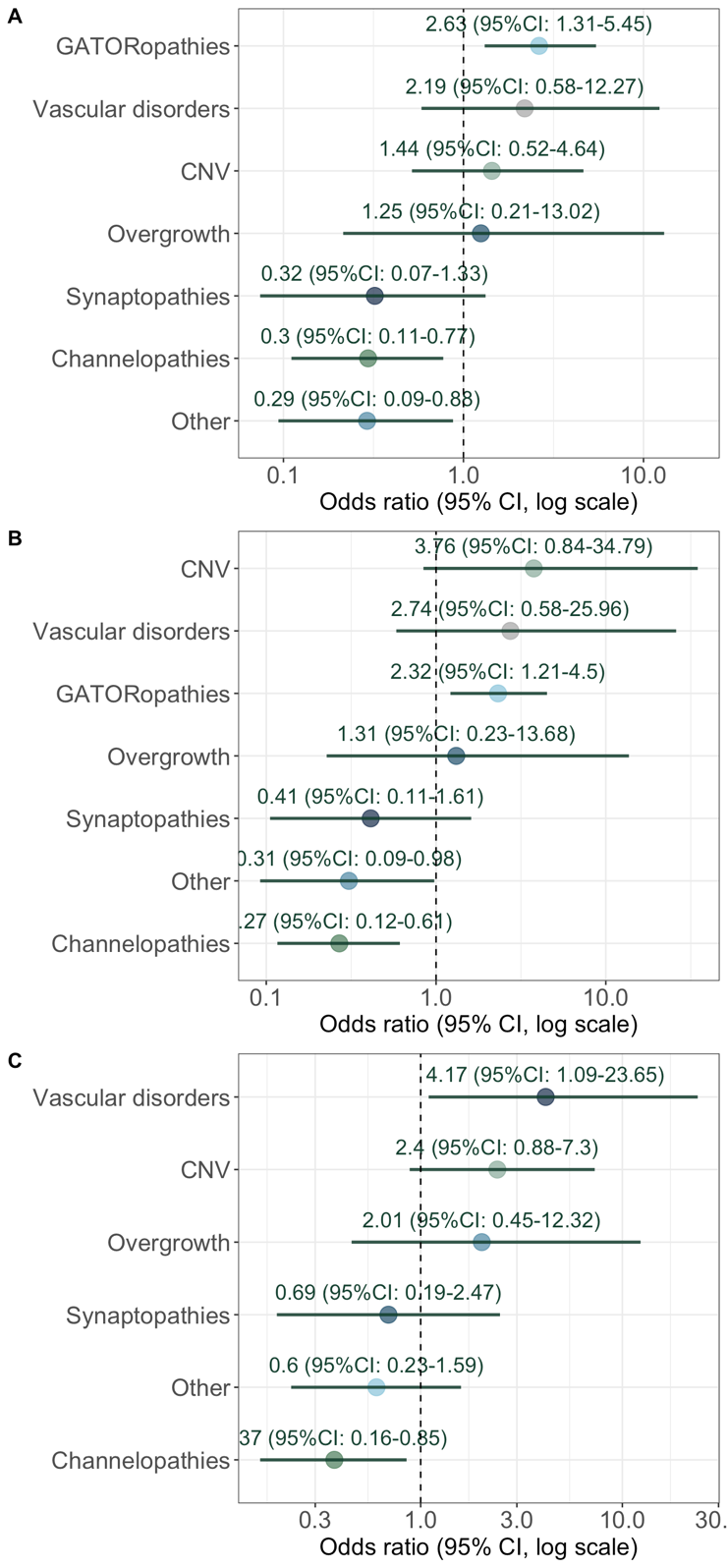

Supplemental Figure 3. Univariate analysis of association of genetic categories with favorable outcome in subcohorts: A) lesional cases only; B) literature cases only; C) all cases besides GATORopathies.

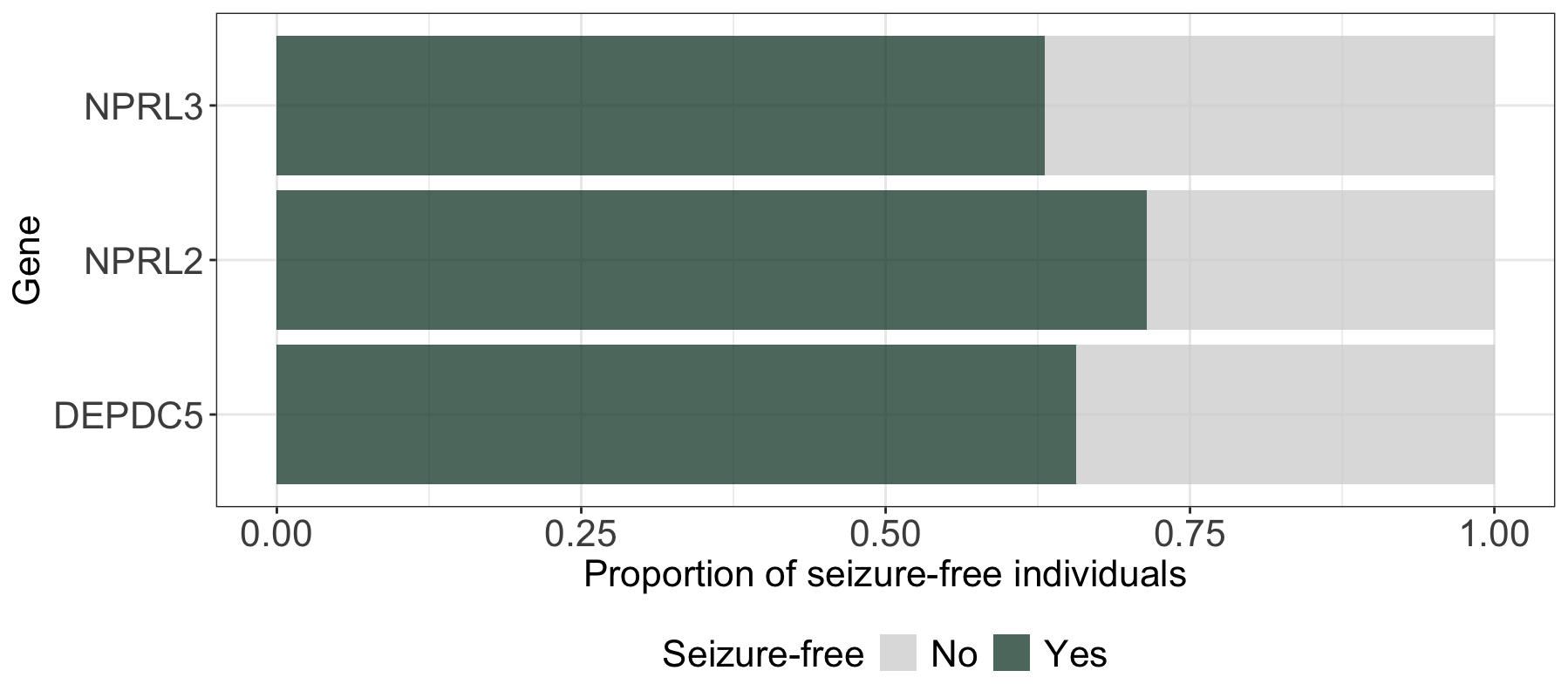

Supplemental Figure 4. Rates of seizure freedom among individuals with GATORopathies. There was no statistically significant difference in the rates of seizure freedom between individuals with variants in *NPRL2*, *NPRL3*, and *DEPDC5*.
